# Machine learning is more accurate and biased than risk scoring tools in the prediction of postoperative atrial fibrillation after cardiac surgery

**DOI:** 10.1101/2024.07.05.24310013

**Authors:** Joyce C Ho, Shalmali Joshi, Eduardo Valverde, Kathryn Wood, Kendra Grubb, Miguel Leal, Vicki Stover Hertzberg

## Abstract

Incidence of postoperative atrial fibrillation (POAF) after cardiac surgery remains high and is associated with adverse patient outcomes. Risk scoring tools have been developed to predict POAF, yet discrimination performance remains moderate. Machine learning (ML) models can achieve better performance but may exhibit performance heterogeneity across race and sex subpopulations. We evaluate 8 risk scoring tools and 6 ML models on a heterogeneous cohort derived from electronic health records. Our results suggest that ML models achieve higher discrimination yet are less fair, especially with respect to race. Our findings highlight the need for building accurate and fair ML models to facilitate consistent and equitable assessment of POAF risk.

## 1 Introduction

Although there have been advancements in cardiac surgery techniques, the incidence of postoperative atrial fibrillation (POAF) following cardiac surgery has not decreased significantly and still ranges from 15% to 50% [1, 2]. Unfortunately, there are short- and long-term adverse outcomes associated with POAF including morbidity, mortality, and longer, more expensive hospitalizations [3, 4, 5, 6, 7]. Early identification of patients at risk for developing POAF has long been desired to guide preventative and treatment strategies. To this end, more than a dozen POAF risk scoring algorithms have been introduced encompassing a variety of risk factors including patient demographics and clinical characteristics as well as surgical characteristics. Yet a recent review found only patient age had no conflicting evidence across existing studies [8]. Moreover, these scoring systems offer moderate discrimination with area under the receiver operating characteristic curve (AUROC) scores ranging between 0.55 and 0.87 and may not generalize broadly as the performance is assessed on relatively small, homogeneous patient populations.

Machine learning (ML) has been proposed as an alternative to achieve better predictive performance [9]. A recent scoping review found that support vector machines (SVM), gradient boosting machines (GBM), and random forests (RF) using clinical characteristics can predict POAF risk more accurately than existing risk scores with promising specificity, sensitivity, and AUROC scores [9]. Three existing works compared multiple ML algorithms with Lu et al [10] and Parise et al. [11] concluding that SVM achieved the best performance while GBM performed the best in Karri et al. [12]. Despite their promise, indiscriminate application of ML models can exacerbate existing health disparities if they are not trained on a representative sample [13].

Unfortunately, significant race and sex disparities exist as the number of patients undergoing cardiac surgery procedures and the outcomes for these patients [14]. Incidence of POAF after coronary artery bypass graft (CABG) surgery is higher in White patients [15]. It has also been suggested males are more likely to experience POAF following CABG [16, 17] although there exists conflicting evidence [18]. However, only 2 studies utilizing ML report the ethnicity composition of the underlying dataset and both studies assessed the performance in populations with less than 4% Black patients [12, 19]. Thus, a crucial unanswered question is whether the better performance of ML algorithms may exacerbate existing disparities.

The objective of this study is to assess both the predictive performance and fairness of existing POAF risk scoring tools with popular ML algorithms on a heterogeneous population, with more than 20% of the patients identifying as Black. We assess the fairness of the predictive models in both race and sex subpopulations. We also restrict our evaluation to common structured data found within electronic health records (EHRs) as such algorithms can provide quicker (and hopefully more accurate) management strategies [9].

## 2 Methods

### 2.1 Data Source

Our study was conducted using de-identified EHRs from the Emory Healthcare clinical data warehouse. Secondary data analysis was approved by the Emory University Institutional Review Board. Adult patients who received cardiac surgery in the outpatient or inpatient setting between January 1, 2013 and December 31, 2017 were included. Cardiac surgery was defined using the Current Procedural Terminology (CPT) codes as either venous grafting for CABG or surgical procedures on cardiac valves (see Supplemental Material for full list). For security purposes, patient identifiers were omitted and certain records were excluded based on the date shifting logic. Patients who had a prior history of atrial fibrillation (AF), defined by the International Classification of Diseases codes of ‘427.31’ for the 9th revision (ICD-9) or ‘I48.XX’ for the 10th revision (ICD-10) were excluded from the study. We used the presence of the AF ICD-9 or ICD-10 code following the cardiac surgery procedure date to identify cases of POAF. The value of 0 was assigned to patients that did not experience POAF and had at least 1 encounter after the cardiac surgery.

All the clinical variables including age, sex, race, height, weight, and blood pressure were extracted from the EHR. We used the most recent value collected within the 1 year prior to the cardiac surgery date. The presence of clinical comorbidities for the risk scoring systems was determined using diagnostic (ICD-9 or ICD-10), procedural (CPT), and medication codes. For the ML clinical variables, we grouped the diagnostic codes using the single-level Clinical Classifications Software (CCS) system and medication codes using Anatomical Therapeutic Chemical (ATC) Level 3 classification codes.

### 2.2 Risk Scores

We evaluated POAF risk scoring systems and incident AF risk scoring systems that utilize commonly collected measures in structured EHR data. Although a recent review identified at least 12 distinct POAF scoring systems, [8] several used echocardiographic measurements such as left atrial dilation, left atrial diameter, and left ventricular ejection fraction which are often captured in unstructured text and are not easily accessible broadly. As such, we focused on the following 8 risk scores: (1) CHADS_2_,[20] (2) CHA_2_DS_2_-VASc,[20] (3) HATCH, [21] (4) COM-AF, [22] (5) C_2_HEST, [23] (6) mC_2_HEST, [24] (7) AFRI [25], and (8) CHARGE-AF [26]. The Python code for the scoring systems is openly available as a GitHub repository (https://github.com/joyceho/afib). The predictor variables for each model can be found in Supplemental Table 3.

### 2.3 Machine Learning Models

Six commonly used ML algorithms were explored that have been previously benchmarked from previous existing studies: (1) logistic regression (LR), (2) decision tree (DT), (3) SVM, (4) RF, (5) GBM, and (6) multi-layer perceptron (MLP). The ML models were constructed using the popular Python open-source software library, `scikit-learn` version 1.5.0 [27]. The ML models were supplied with age, race, gender, CCS, and ATC codes. CCS and ATC codes that were not present in at least 20% of the patients were excluded. A total of 71 variables were supplied as input to the models. Exhaustive hyperparameter optimization was performed using 5-fold cross validation on the training dataset (see Supplemental Table 5 for the parameter search space for each model). The optimal hyperparameter for each model was identified using AUROC.

### 2.4 Training and Evaluation

We used stratified Monte Carlo cross validation to randomly split the data into 70-30% train-test. This process was repeated 10 times to assess model performance. Data imputation was required for age, height, weight, and blood pressure. Mean imputation from the training data was used. Predictive performance was measured using AUROC and area under the precision recall curve (AUPRC) on the test set. AUROC and AUPRC were calculated using the scikit-learn package on the test data. We also assessed the fairness of the models on the sex (female and male) and race (White and Black/Other) subgroups. Two popular group fairness metrics were used, demographic parity ratio (DPR) and equalized odds ratio (EQR). DPR measures whether the predictive proportion of POAF across the subgroups are equal (i.e., the prediction risk should be independent of sex or race). EQR ensures the true positive rate and false positive rate of predictions are the same across the subgroups. Both DPR and EQR range from 0 to 1 with 1 indicates fairness across the subgroups. DPR and EQR were computed using the `fairlearn` package version 0.10.0.[28] All analyses were performed using Python version 3.9.7.

## 3 Results

### 3.1 Patient Characteristics

Out of the final study population of 4961 patients, 1953 (39.4%) experienced POAF following cardiac surgery with an average onset of xxx. Baseline characteristics of the overall study population and the 2 outcome groups (no POAF and POAF) are reported in Table 1. The incidence of POAF experienced in males (40.1%) and Whites (42.5%) was statistically higher than in females (37.9%) and Blacks (32.3%), respectively.

**Table 1:** Baseline characteristics in patients with and without POAF.

|  | Missing | Overall<br>(n=4961) | No POAF<br>(n=3008) | POAF<br>(n=1953) | P-Value |
| --- | --- | --- | --- | --- | --- |
| Age, mean (SD) | 19 | 64.6 (12.4) | 63.3 (13.1) | 66.5 (10.9) | <0.001 |
| Male, n (%) | 0 | 3300 (66.5) | 1976 (65.7) | 1324 (67.8) | 0.133 |
| Ethnicity, n (%) | 0 |  |  |  | <0.001 |
| White |  | 3426 (69.1) | 1970 (65.5) | 1456 (74.6) |  |
| Black |  | 1085 (21.9) | 735 (24.4) | 350 (17.9) |  |
| Other |  | 450 (9.1) | 303 (10.1) | 147 (7.5) |  |
| CHF, n (%) | 0 | 2066 (41.6) | 1226 (40.8) | 840 (43.0) | 0.123 |
| HTN, n (%) | 0 | 2999 (60.5) | 1888 (62.8) | 1111 (56.9) | <0.001 |
| DM, n (%) | 0 | 1382 (27.9) | 864 (28.7) | 518 (26.5) | 0.098 |
| Stroke, n (%) | 0 | 959 (19.3) | 560 (18.6) | 399 (20.4) | 0.123 |
| Vascular Disease, n (%) | 0 | 2004 (40.4) | 1253 (41.7) | 751 (38.5) | 0.027 |
| COPD, n (%) | 0 | 509 (10.3) | 307 (10.2) | 202 (10.3) | 0.914 |
| CAD, n (%) | 0 | 4041 (81.5) | 2503 (83.2) | 1538 (78.8) | <0.001 |
| SHF, n (%) | 0 | 379 (7.6) | 228 (7.6) | 151 (7.7) | 0.887 |
| Hyperthyroid, n (%) | 0 | 18 (0.4) | 14 (0.5) | 4 (0.2) | 0.211 |
| Valvular, n (%) | 0 | 3099 (62.5) | 1783 (59.3) | 1316 (67.4) | <0.001 |
| PVD, n (%) | 0 | 1265 (25.5) | 723 (24.0) | 542 (27.8) | 0.004 |
| Obesity, n (%) | 0 | 420 (8.5) | 255 (8.5) | 165 (8.4) | 1.000 |
| Height (m), mean (SD) | 65 | 67.7 (4.6) | 67.4 (4.6) | 68.0 (4.5) | <0.001 |
| Weight (kg) , mean (SD) | 41 | 192.0 (45.8) | 189.9 (44.5) | 195.3 (47.5) | <0.001 |
| SBP, mean (SD) | 40 | 135.3 (22.3) | 135.1 (22.2) | 135.5 (22.4) | 0.559 |
| DBP, mean (SD) | 41 | 74.4 (12.9) | 74.8 (12.9) | 73.8 (12.8) | 0.015 |

### 3.2. Performance Comparison

Table 2 summarizes the discrimination and fairness performance of the 14 models (8 risk scoring algorithms and 6 ML models). For each performance metric, the value represents the mean across the 10 test splits. Statistical significance in discrimination performance between any 2 models was assessed using a one-tailed paired t-test that the difference is greater than 0 (i.e., one model consistently outperforms the other).

**Table 2:** Average discrimination and fairness performance of the prediction models across 10 Monte Carlo cross-validation splits.

| Model | AUROC | AUPRC | Race |  | Sex |  |
| --- | --- | --- | --- | --- | --- | --- |
|  |  |  | DPR | EQR | DPR | EQR |
| RF | 0.671 | 0.558 | 0.824 | 0.816 | 0.967 | 0.946 |
| GBM | 0.666 | 0.553 | 0.837 | 0.835 | 0.975 | 0.948 |
| SVM | 0.658 | 0.540 | 0.828 | 0.827 | 0.976 | 0.958 |
| LR | 0.645 | 0.521 | 0.659 | 0.618 | 0.949 | 0.903 |
| DT | 0.630 | 0.534 | 0.970 | 0.950 | 0.962 | 0.942 |
| MLP | 0.622 | 0.494 | 0.789 | 0.772 | 0.953 | 0.923 |
| CHARGE-AF | 0.578 | 0.443 | 0.896 | 0.895 | 0.941 | 0.926 |
| AFRI | 0.568 | 0.474 | 1.000 | 1.000 | 1.000 | 1.000 |
| COM-AF | 0.550 | 0.416 | 0.993 | 0.980 | 0.884 | 0.858 |
| HAVOC | 0.527 | 0.401 | 0.999 | 0.999 | 0.999 | 0.998 |
| HATCH | 0.527 | 0.400 | 1.000 | 1.000 | 1.000 | 1.000 |
| mC<sub>2</sub>HES<sub>T</sub> | 0.526 | 0.408 | 1.000 | 1.000 | 1.000 | 1.000 |
| CHA<sub>2</sub>DS<sub>2</sub>-VASc | 0.525 | 0.408 | 1.000 | 1.000 | 1.000 | 1.000 |
| CHADS<sub>2</sub> | 0.517 | 0.401 | 1.000 | 1.000 | 1.000 | 1.000 |
| C<sub>2</sub>HES<sub>T</sub> | 0.509 | 0.401 | 1.000 | 1.000 | 1.000 | 1.000 |

The ML model that achieved the best discrimination was RF with AUROC and AUPRC of 0.671 and 0.558, respectively. Only GBM yielded a p-value above 0.001 for AUROC (0.03) and AUPRC (0.06) during the one-tailed paired t-test between RF and the other 5 ML models. Among the risk scoring systems, CHARGE-AF achieved the best performance with AUROC and AUPRC of 0.585 and 0.449, respectively. Notably, all 6 ML models outperformed CHARGE-AF and the other risk scoring tools at statistically significant levels (p-value < 0.001) for both discrimination metrics.

The risk scoring systems generally yielded the best group fairness concerning race as all but CHARGE-AF, COM-AF, and HAVOC resulted in both DPR and EQR of 1. Notably, CHARGE-AF is the only risk scoring system incorporating race as a variable (see Supplemental Table 3), yet achieves the worst group fairness. In contrast, all the ML models except DT perform worse in terms of DPR and EQR to CHARGE-AF (DPR = 0.896 and EQR = 0.895).

A similar group fairness trend is observed for sex in terms of risk scoring systems again as CHARGE-AF, COM-AF, and HAVOC do not yield DPR and EQR of 1. However, the ML models achieve slightly better performance than CHARGE-AF and COM-AF in terms of DPR and EQR for race. Surprisingly, COM-AF which incorporates sex as a variable yields the worst DPR and EQR performance with values of 0.884 and 0.858, respectively.

## 4 Discussion

In this study, we evaluated the performance of 6 ML models and 8 risk scoring algorithms to predict POAF. We demonstrated that RF outperformed the other ML methods and all of the risk scores considered in terms of AUROC and AUPRC. Furthermore, there were statistical differences between the discrimination performance of RF and the other models except for the GBM algorithm. The AUROC and AUPRC of these 14 models were all under 0.671 and 0.558 respectively. Compared to the existing ML studies, the discrimination performance is lower as they achieved an AUROC of at least 0.72. However, these models used indicators related to cardiac surgery which are not commonly available in the structured EHR data.

In contrast to the discrimination performance, six of the risk scores outperformed all of the ML methods and three of the risk scores with respect to metrics of fairness. In fact, the results indicate the ML models exacerbated race and sex differentials when used for POAF prediction, which is consistent with existing evidence for other outcomes such as cardiovascular risk [29], dermatology [30], and population health [31]. Thus, better discrimination performance may not always be desired as it might exacerbate existing race and sex disparities. This suggests further investigation is necessary to holistically assess the efficacy of ML algorithms for POAF prognostication in real clinical contexts, [32] and whether bias mitigation mechanisms should be adopted to minimize disparities in outcomes and interventions.

## Data Availability

Data provided in the present study are not available but the code used is available.

## Abbreviations

POAF: post-operative atrial fibrillation
AUROC: area under the receiver operating characteristic curve
ML: machine learning
SVM: support vector machines
GBM: gradient boosting machines
RF: random forests
CABG: coronary artery bypass graft

## 5 Supplemental Information

### 5.1 Risk scoring algorithms

The variables used for each of the 8 risk scores are summarized in Table 3. While other risk scoring systems have been developed using POAF as an event of interest[8], they are not benchmarked in our study as they use variables such as left atrial dilatation, left atrial diameter, left ventricular ejection fraction, and length of stenosis. The scores in Table 3 can be computed from demographic information, diagnosis tables (ICD-9/ICD-10), vital signs commonly collected, and medication tables.

**Table 3:**
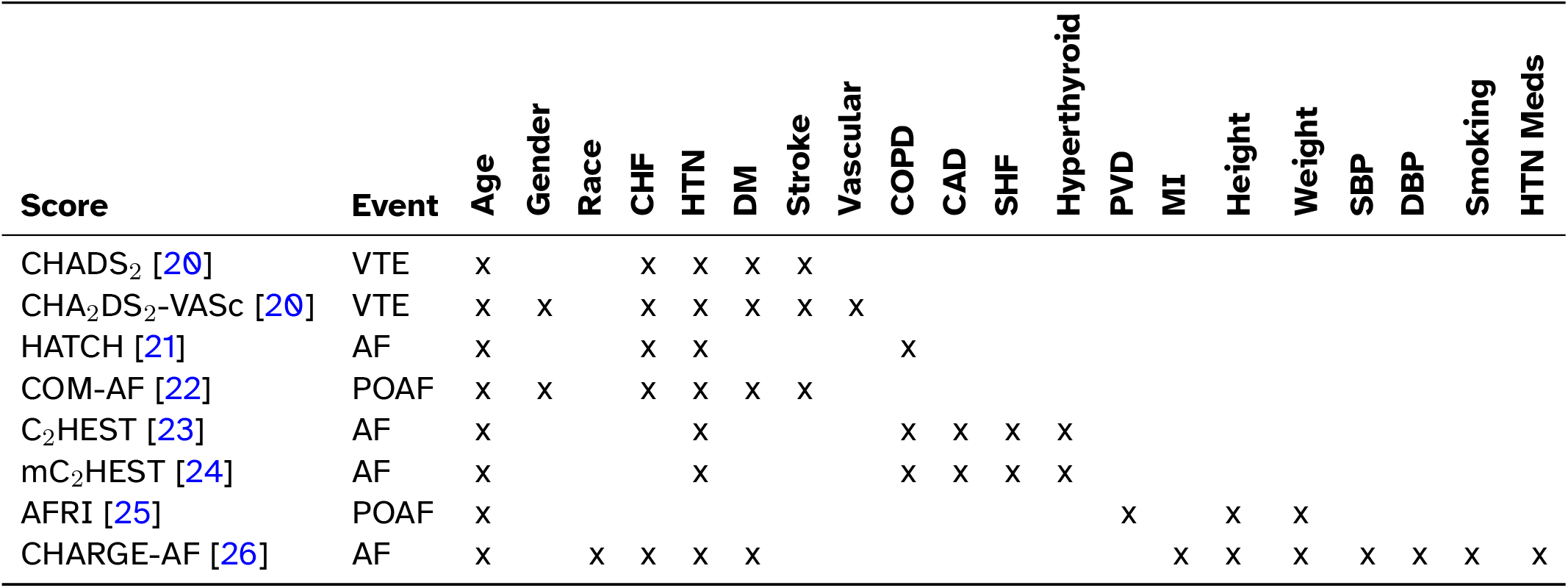
Risk scoring systems, the original event of interests, and their associated variables. The events of interest are thromboembolic events in patients with atrial fibrillation (VTE), incident AF (AF), and POAF.

### 5.2 ML for POAF

A recent scoping review identified 7 papers that used ML for predicting POAF after cardiac surgery.[9] Of the 7 studies, 3 relied on electrocardiogram data while the remaining 4 used clinical documentation, administrative data, or Holter monitoring. The sample size, ethnicity composition, and model performance of the 4 ML studies using administrative data are summarized in Table 4. As can be seen, none of the patient populations contains more than 3.4% Black.

**Table 4:**
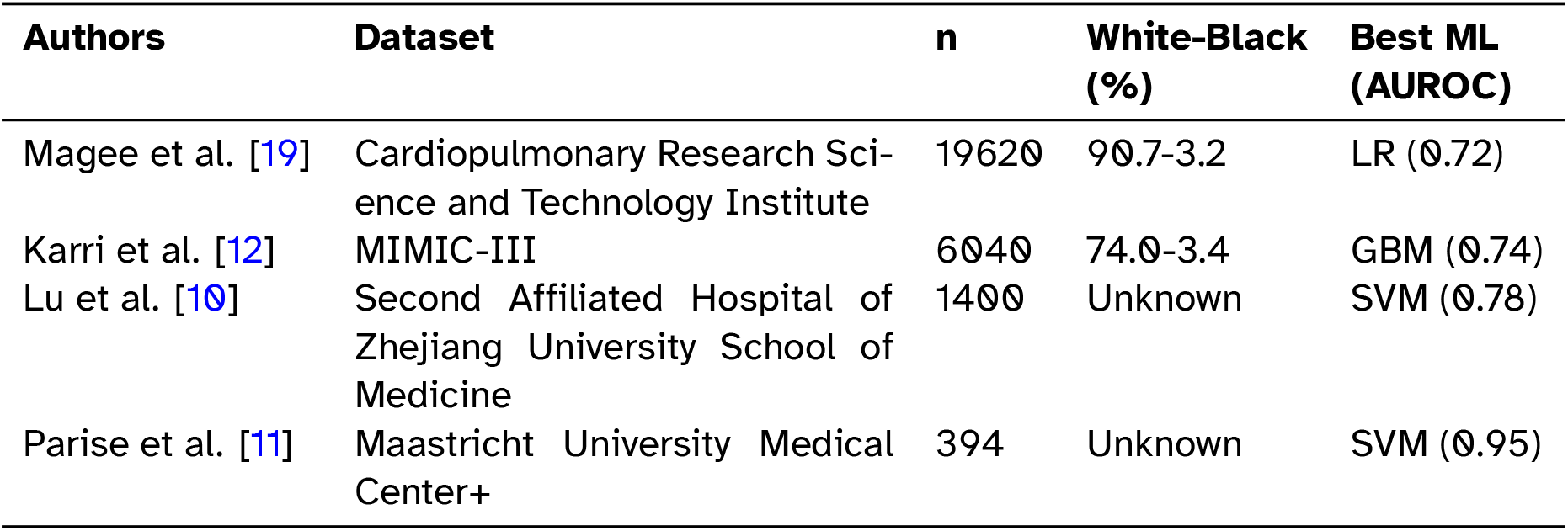
Previous ML studies for POAF prediction.

Table 5 summarizes the hyperparameter search space for each of the ML models. For each train-test split and ML model, GridSearchCV in scikit-learn was performed using 5-folds on the train split to find the optimal hyperparameter values. The ML model is then retrained using the optimal hyperparameter values and the performance is evaluated on the test set.

**Table 5:**
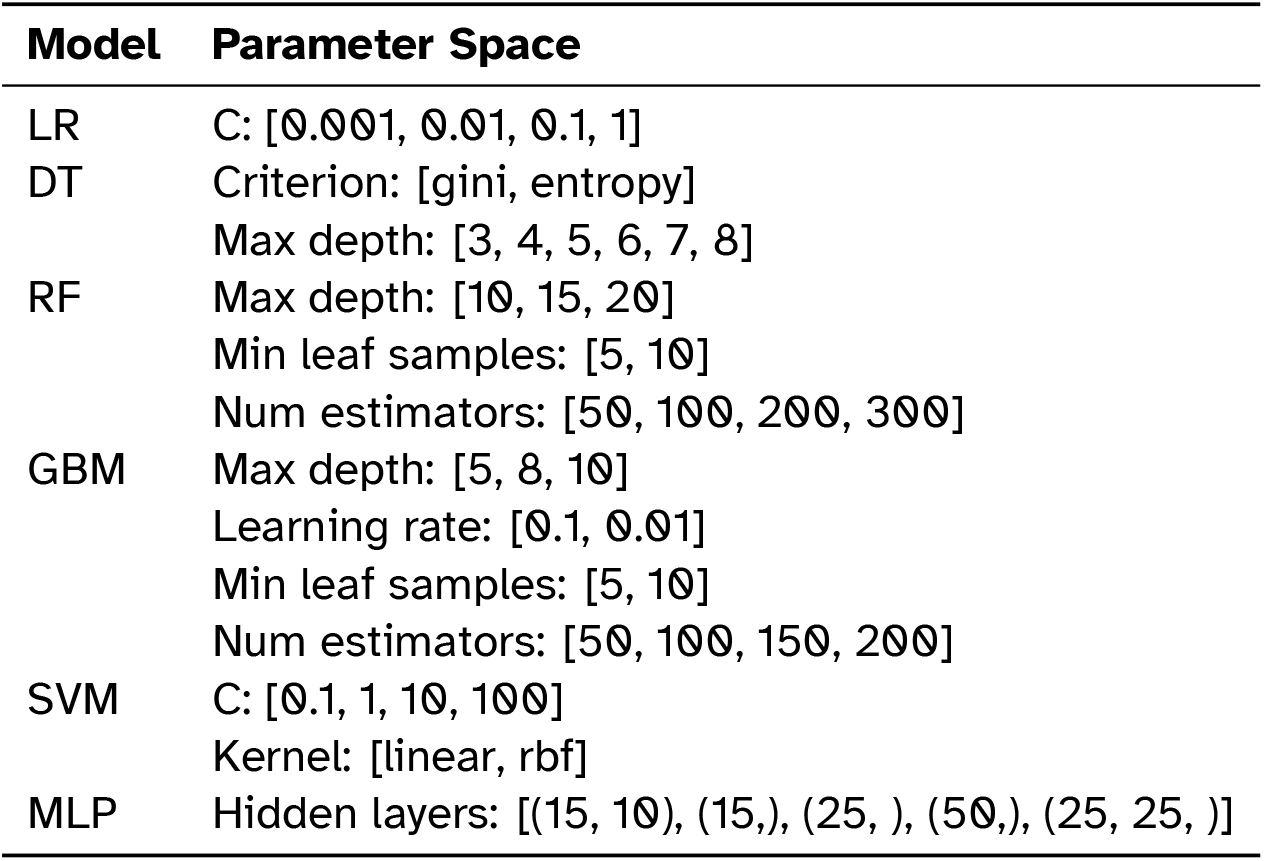
Hyperparameter search space for the different ML models.

